# Pre-Existing Mood Disorder is Associated with Higher Odds of Withdrawal of Life-Sustaining Treatment (WOLST) After Severe Stroke: A Retrospective Cohort Study

**DOI:** 10.64898/2026.09.25.26363539

**Authors:** Claire G. McHugh, Christian M. Kabongo, Matthew Siegel, Lester Y. Leung

**Affiliations:** Tufts University School of Medicine, Boston, MA USA; Tufts Medical Center, Department of Neurology, Boston, MA USA

**Keywords:** Acute Ischemic Stroke, Intracerebral Hemorrhage, Intraparenchymal Hemorrhage, Mood Disorders, Stroke, Withdrawal of Life-Sustaining Treatment

## Abstract

**Background/Aims:** Pre-stroke depression has been shown to be associated with worse functional outcomes and depression after stroke. However, it is not currently well understood whether pre-stroke mood disorders (e.g., depression, anxiety, mood and psychotic disorders, substance use disorder) are associated with withdrawal of life-sustaining treatment (WOLST) after a severe stroke.

**Methods:** This was a retrospective cohort study within the Tufts Vascular Neurology Registry of patients hospitalized with severe acute ischemic stroke (AIS, defined as NIHSS *≥*10) or severe nontraumatic intracerebral hemorrhage (ICH, defined as ICH score *≥*2) between 2018 and 2022. Multivariate logistic regression was performed to identify predictors of WOLST.

**Results:** This study included 405 patients with severe stroke. Patients who opted for WOLST were more likely to be older, female, have AIS, and experience more severe strokes. In an adjusted analysis (basic demographics, stroke type), a pre-stroke mood disorder was associated with WOLST (OR 2.15, 95% CI 1.11-4.15). In models stratified by stroke type (AIS vs. ICH) and adjusted for stroke severity, WOLST was associated with age in both groups and with mood disorder in patients with ICH. The WOLST event rate was higher in the ICH group (19% vs 11%).

**Conclusion:** Pre-stroke mood disorder was associated with increased rates of withdrawal of care after severe stroke in this patient population. Opportunities to actively treat depression may be useful to include in goals of care discussions to mitigate the potential impact of this factor in decision-making. Consistent with prior studies, we observed a higher proportion of WOLST in older, female, and ICH patients. Future studies regarding the quality and content of goals of care discussions may help elucidate the differing effects of pre-stroke mood disorders on WOLST by stroke type seen in this study.

## Introduction

Withdrawal of life-sustaining treatment (WOLST) commonly occurs in patients with severe stroke and is one of the most consequential decisions affecting end-of-life stroke care.^1,2^ These decisions require consideration or prognosis, anticipated functional and cognitive outcomes, patient preferences, and shared decision-making between clinicians, patients, and their families.^3-5^ Several clinical and demographic factors are have been associated with WOLST, including older age and pre-existing comorbidities such as cardiomyopathy, dementia, prior stroke/TIA, or metastatic cancer.^6-8^ Psychosocial factors and perceptions of post-stroke quality of life may also influence treatment decisions, particularly when substantial cognitive disability is anticipated.^8-10^ Despite recognition of these factors, the influence of pre-existing mental health conditions on WOLST decisions remains poorly understood. Depression and other mood disorders are associated with stroke outcomes, and pre-existing depressive symptoms have been independently associated with short-term and in-hospital mortality after stroke.^11,12^ However, whether pre-existing mood disorders are associated with WOLST after severe stroke has not been rigorously studied. Therefore, we sought to evaluate the association between pre-stroke mood disorders and WOLST among patients with severe stroke.

## Methods

### Patient Population

The study population included patients age 18 and older hospitalized with a severe acute ischemic stroke (AIS) or severe nontraumatic intraparenchymal hemorrhage (IPH) at Tufts Medical Center between 2018 and 2022 within the framework of the Tufts Medical Center Vascular Neurology Registry. Severe stroke was defined as AIS with National Institutes of Health Stroke Scale (NIHSS) *≥* 10 and IPH with Intracerebral Hemorrhage (ICH) Score *≥* 2.^13,14^ This study was approved by the Tufts Institutional Review Board (#12151).

### Predictors and Outcomes

The electronic medical record was used to record study variables including age, sex, race/ethnicity, treatment modalities (intravenous thrombolysis [IVT] and endovascular thrombectomy), the patient’s WOLST decision, and history of mood disorders (psychotic disorders, depression, anxiety, bipolar disorder, post-traumatic stress disorder). Patients were placed in the mood disorder group if they met at least one of the following criteria in the year preceding their stroke: a mood disorder documented in the problem list, admission note, outpatient note, or a prescription for antidepressants, antipsychotics, mood stabilizers, or anti-anxiety medications associated with an encounter reporting mood symptoms or a psychiatric condition that is being treated.

### Statistical Analysis

Statistical analyses in this study were performed in Stata (v18.5, College Station, Texas). First, univariate t-tests and chi-square analysis were conducted to evaluate differences in patient characteristics across mood disorder groups. Then, multivariable logistic regression (guided by clinician topical expertise) were conducted to identify independent predictors of WOLST. The primary multivariable logistic regression model evaluated the likelihood of WOLST using the entire study population. WOLST was defined as a binary of withdrawal of life-sustaining treatment or not. The model was adjusted for age, sex, race/ethnicity, and stroke type (AIS or ICH). The exposure variable was a previous mood disorder measured as a binary. Stratified analyses (by stroke type, AIS and IPH) were also conducted, including stroke type-specific severity and treatment variables (NIHSS, IVT, and EVT for AIS; ICH score for IPH).

## Results

### Study Cohort and WOLST Prevalence

From the Tufts Medical Center Vascular Neurology Registry, 405 patients met the inclusion criteria between 2018 and 2022 (**Table 1**). Of these patients, 52 (12.8%) underwent withdrawal of life-sustaining treatment (WOLST).

**Table 1.** Demographic and Clinical Characteristics of Study Population. *\* P < 0*.*05. * = statistically significant data. SD = standard deviation, ICH score = intracerebral hemorrhage score, NIHSS = National Institutes of Health Stroke Scale, and EVT = endovascular thrombectomy*

| Characteristic | No WOLST | WOLST | Total | P-Value |
| --- | --- | --- | --- | --- |
| N (%) | 353 (87.2%) | 52 (12.8%) | 405 |  |
| History of Mood Disorder |  |  |  |  |
| No (%) | 239 (67.7%) | 30 (57.7%) | 269 | 0.153 |
| Yes (%) | 114 (32.3%) | 22 (42.3%) | 136 |  |
| Age, years at time of episode |  |  |  |  |
| (SD) | 68.39 (16.02) | 80.83 (12.93) | 69.99 (16.19) | <0.001* |
| Sex |  |  |  |  |
| Female (%) | 180 (82.9%) | 37 (17.1%) | 217 | 0.006* |
| Male (%) | 173 (92.0%) | 15 (8.0%) | 188 |  |
| Race/Ethnicity |  |  |  |  |
| Non-Hispanic White (%) | 259 (87.2%) | 38 (12.8%) | 297 | 0.855 |
| Non-Hispanic Black (%) | 34 (89.5%) | 4 (10.5%) | 38 |  |
| Non-Hispanic Asian (%) | 60 (85.7%) | 10 (14.3%) | 70 |  |
| Mean GINI Index by Zip |  |  |  |  |
| Code (SD) | 0.44 (0.06) | 0.46 (0.08) | 0.45 (0.07) | 0.044* |
| Primary Cerebrovascular |  |  |  |  |
| Diagnosis |  |  |  |  |
| Acute ischemic stroke (%) | 282 (89.0%) | 35 (11.0%) | 317 | 0.040* |
| Intraparenchymal hemorrhage (%) | 71 (80.7%) | 17 (19.3%) | 88 |  |
| NIHSS (SD) | 16.87 (5.5) | 19.45 (5.6) | 17.19 (5.55) | 0.005* |
| ICH Score (SD) | 2.40 (0.91) | 3.07 (0.70) | 2.52 (0.91) | 0.009* |
| tPA |  |  |  |  |
| No tPA (%) | 244 (86.5%) | 38 (13.5%) | 282 | 0.563 |
| Yes tPA (%) | 109 (88.6%) | 14 (11.4%) | 123 |  |
| EVT |  |  |  |  |
| No EVT (%) | 210 (86.1%) | 34 (13.9%) | 244 | 0.470 |
| Yes EVT (%) | 132 (88.6%) | 17 (11.4%) | 149 |  |

### Univariate analyses

WOLST occurred more frequently among patients with IPH (19.3%) compared to those with AIS (11.0%). Patients who underwent WOLST were significantly older (80.8 vs. 68.4 years, p < 0.001), more commonly female (17.1% vs. 8.0% among males, p = 0.006), and resided in areas with greater income inequality as measured by the GINI index (0.46 vs. 0.44, p = 0.044). They also had greater stroke severity, as evidenced by higher NIHSS scores (19.45 vs. 16.87, p = 0.005) and ICH scores (3.07 vs. 2.40, p = 0.009). Rates of thrombolysis and endovascular thrombectomy did not differ significantly between groups. Pre-existing mood disorders were more common among patients who underwent WOLST (42.3% vs. 32.3%), although this difference was not statistically significant on univariate analysis (p = 0.153).

### Associations of patient characteristics with WOLST

In multivariable logistic regression analysis, increasing age was independently associated with WOLST (OR 1.07, p < 0.001) (**Table 2**). Pre-existing mood disorder was also independently associated with WOLST (OR 2.10, p < 0.027). In the adjusted model including demographic and socioeconomic covariates, mood disorder remained significantly associated with WOLST (OR 2.15, 95% CI 1.11–4.15).

**Table 2.**
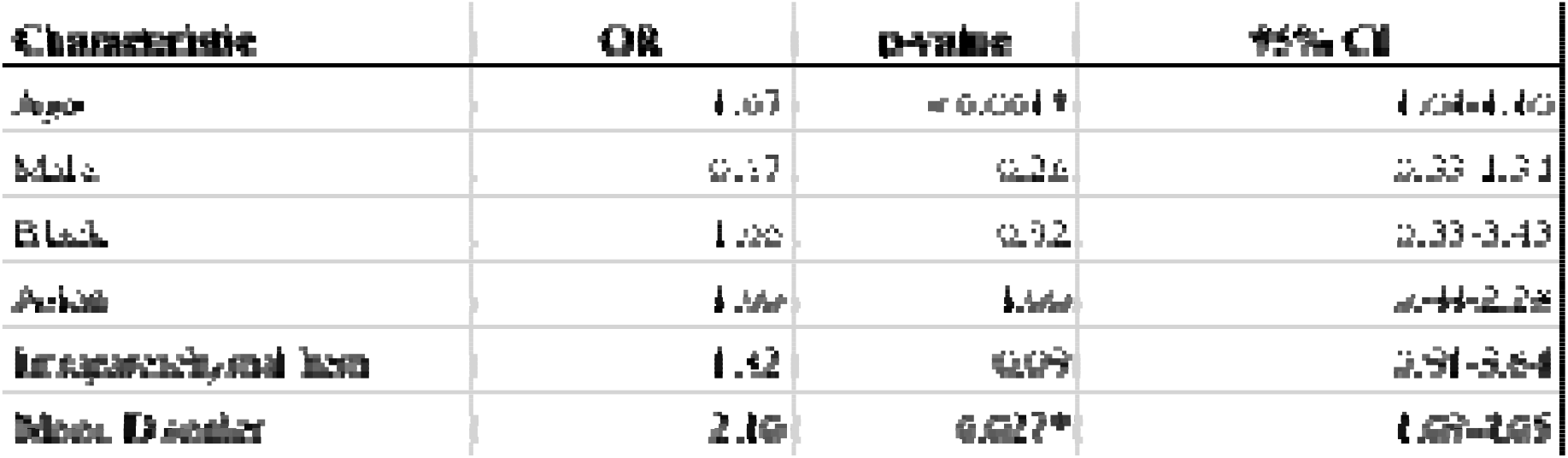
Associations of patient characteristics and withdrawal of life-sustaining treatment (WOLST). *\* P < 0*.*05. * = statistically significant data. OR = odds ratio, CI = confidence interval, and hem. = hemorrhage*.

### Stratified analysis by stroke subtype

When stratified by stroke subtype, increasing age remained associated with WOLST in both acute ischemic stroke and intracerebral hemorrhage cohorts (AIS: OR 1.06, p < 0.001; ICH: OR 1.11, p < 0.014) (**Table 3**). Pre-existing mood disorder was significantly associated with WOLST among patients with intracerebral hemorrhage (OR 4.88, p < 0.046) but not among those with acute ischemic stroke (OR 1.45, 95% CI 0.69–3.41).

**Table 3.**
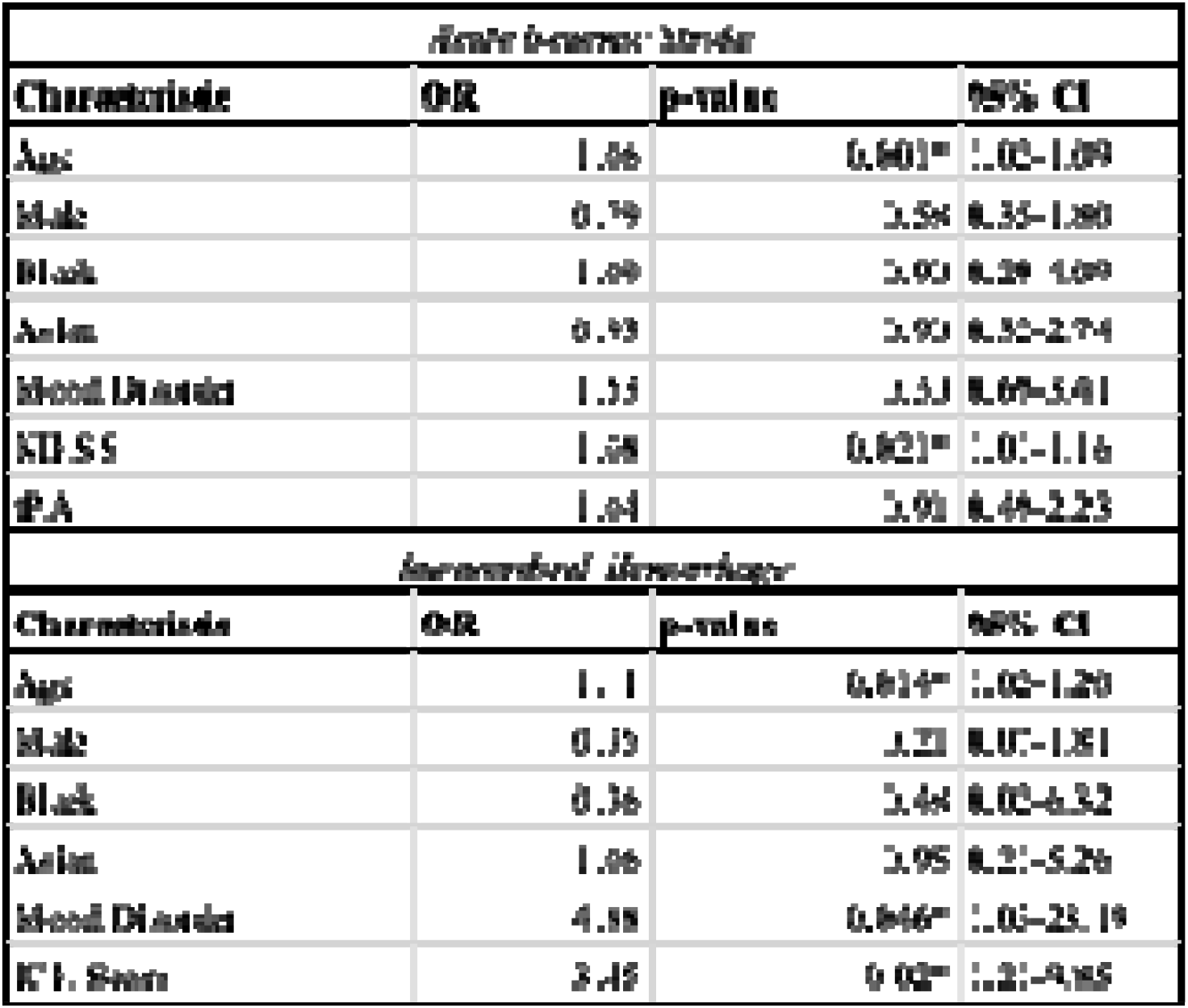
Analysis Stratified by Stroke Type (Acute Ischemic Stroke and Intraparenchymal Hemorrhage). *\* P < 0*.*05. * = statistically significant data. OR = odds ratio, CI = confidence interval, ICH score = intracerebral hemorrhage score, and NIHSS = National Institutes of Health Stroke Scale*.

## Discussion

In this retrospective cohort study consisting of patients who were hospitalized with severe stroke, we discovered that pre-existing mood disorders are independently linked to increased odds of withdrawal of life-sustaining treatment, even after adjustment for demographic and socioeconomic factors. Increasing age was also strongly associated with WOLST; more specifically, WOLST increased by 7% for each additional year, and patients with IPH had higher WOLST rates than those with AIS, findings that align with previous studies on severe stroke and end-of-life decision-making.^8^ While prior literature has primarily focused on clinical severity, demographic factors, and prognosis as contributors to WOLST decisions, our findings suggest that pre-existing mood disorders may also influence goals-of-care decisions after stroke. These findings indicate that the severity of neurologic injury, in conjunction with pre-stroke characteristics such as psychosocial status, is an important consideration in informing end-of-life decision-making.

Interestingly, the association between pre-existing mood disorder and WOLST appeared to be driven primarily by patients with IPH, as the association disappeared when the analysis was focused on patients with AIS. The underlying reasons that explain this distinction between IPH and AIS likely include biological, clinician behavior, and prognostic factors. Patients with IPH typically have profound neurological deficits and a more prolonged recovery timeline compared to patients with AIS. ^15,16^ For patients with AIS, there may be more optimism surrounding their clinical outcome, which may be rooted in the difference in treatment availability, as reperfusion therapy and other acute interventions are often offered to AIS patients.^15-18^ These differing expectations of the outcome may influence goals-of-care discussions between clinicians and family. Additionally, clinicians systematically overestimate IPH severity relative to AIS, including overestimating the volume of the lesions in IPH.^19^ This, in turn, leads to prognostic pessimism, and if these patients have a history of mood disorder, it can amplify the perceptual bias of the patient’s ability to respond well to rehabilitation.

Beyond the observed relationship between pre-existing mood disorders and WOLST in patients with intracerebral hemorrhage, our analysis also identified associations between WOLST and sex in our cohort. Prior studies have shown that women with stroke are more likely to live alone, be widowed, and have reduced caregiver support.^20,21^ Women have additionally been shown to experience worse functional outcomes after stroke and differences in treatment patterns, including higher rates of palliative care utilization and lower likelihood of receiving thrombolysis or thrombectomy.^20–22^ Consistent with this literature, female patients in our cohort demonstrated a higher probability of WOLST, though this association was not significant in multivariable analyses.

A strength of this study is that it builds upon the established Tufts Vascular Neurology Registry, which has supported multiple prior published analyses examining stroke outcomes and social determinants of health. The registry has enabled exploration of disparities in acute stroke care encompassing patient and hospital characteristics, including EMS utilization, stroke-code activation, and treatment times, as well as examination of neighborhood socioeconomic conditions and their relationship with post-stroke functional recovery.^23,24^ Additionally, the registry has facilitated longitudinal characterization of post-stroke neurologic complications following reperfusion therapy, including late-onset movement disorders and their association with functional outcomes, and detailed characterization of differences in clinical presentation and early neurologic course across age groups.^25,26^ Collectively, these studies demonstrate the registry’s ability to capture detailed patient-level clinical, treatment, socioeconomic, and longitudinal outcome data. Building on this established infrastructure, the use of this registry allowed for detailed individual patient-level characterization. The registry enabled us to study both patients with severe AIS and IPH, strengthening the relevance and robustness of our findings in this high-risk population. The combination of detailed patient-level characterization and representation of both major severe stroke subtypes strengthens the relevance of these findings to understanding factors associated with end-of-life decision-making after severe stroke.

Our study had notable limitations. Identifying pre-existing mood disorders in retrospective studies can be challenging, as psychiatric diagnoses may be underreported or inconsistently documented. We attempted to mitigate this limitation by leveraging a comprehensive electronic medical record system spanning multiple health systems and by incorporating several methods to improve diagnostic capture. Additionally, our study was unable to assess the specific content, quality, or context of goals-of-care discussions because documentation of these conversations is often inconsistent. Future prospective studies examining clinician-family communication and decision-making processes may further clarify the mechanisms underlying WOLST decisions after severe stroke. Finally, while pre-existing mood disorders were associated with higher odds of WOLST, the quality of treatment of these mood disorders could not be consistently ascertained.

## Conclusion

This study establishes that pre-existing mood disorders are independently associated with increased odds of WOLST after severe stroke. This association appeared the strongest amongst patients with IPH, which may be explained by the psychosocial and structural factors relating to care for these patients in combination with the severity of the neurological injury. Given the following, goals of care discussions after severe stroke should include discussion and evaluation of the patient’s mental health history, the social determinants of health that impact them, and the potential prognostic bias. Although psychiatric illness may influence recovery and engagement with rehabilitation, these conditions should not independently predetermine treatment decisions after stroke, underscoring the importance of comprehensive mental health evaluation and support during goals-of-care discussions. This study’s findings can inform future studies analyzing clinician-family communication, decision-making, and prognostication surrounding WOLST to improve supportive services for these patients and make post-stroke decision-making more patient-centered and equitable.

## Data Availability

All data produced in the present work are contained in the manuscript.

